# COVID-19 associated anxiety enhances tinnitus

**DOI:** 10.1101/2020.07.02.20145532

**Authors:** Li Xia, Gang He, Yong Feng, Xiaoxu Yu, Xiaolong Zhao, Zhengnong Chen, Shankai Yin, Jian Wang, Jiangang Fan, Chuan Dong

## Abstract

**Objectives:** To investigate if the anxiety associated with COVID-19 is a promoting factor to tinnitus.

**Methods:** A retrospective research design was used to compare the clinical characteristics of tinnitus between the patients in 2020 under pandemic pressure and those from the matching period in 2019. While anxiety was quantified using the Zung’s Self-rating Anxiety Scale (SAS), tinnitus severity was evaluated using the Tinnitus Handicap Inventory (THI) questionnaire and the test of minimum masking level (MML). The assessments were repeated after the sound therapy plus educational counselling (STEC) and compared with EC alone therapy.

**Results:** A large increase in anxiety was evident in 2020 in both case rate and SAS. The treatment of both methods was less effective in 2020. SAS, THI and MML were all deteriorated after the EC alone treatment in 2020, while an improvement was seen in 2019. This suggests that EC alone could not counteract the stress by COVID-19 at all, and the stress, if not managed well, can significantly increase the severity of tinnitus and associated anxiety.

**Conclusions:** By using the EC subgroup in virtual control, we conclude that anxiety can serve as a promoting factor to tinnitus. We believe that this is the first study report that confirm the causative/promotive role of anxiety on tinnitus.

## Introduction

The spread of coronavirus disease 2019 (COVID-19) has already reached pandemic proportions, affecting the majority of countries, areas, and territories across the world (Remuzzi et al., 2020). By the end of June 2020, over nine million people had tested positive for COVID-19 with the death toll increasing to more than 484,000 globally (World Health Organization, 2020). Decisive containment measures in China have reduced new cases and the spread of infection (Liu et al., 2020). However, worries about the spread of the disease, living difficulties, and financial burden related to the pandemic are likely to have had negative psychosocial impacts on residents, as reported by many recent studies (Brooks et al., 2020; Lu et al., 2020; Wang et al., 2020b). It would be reasonable, therefore, to expect an increase in the incidence of disorders that are associated with psychological issues.

Tinnitus is typically referred to as the perception of sound in the absence of an acoustic stimulus or that is only generated by structures in the ear, commonly described as ringing in one or both ears (Bauer, 2018). While the exact mechanisms of tinnitus remain unclear, many risk or promoting factors have been identified, including sensorineural hearing loss, vestibular schwannoma, ototoxic medications, and emotional stress (Baguley et al., 2013). Tinnitus has been linked to stress and related disorders in many previous studies. This link has been thoroughly reviewed, repeatedly, by different authors (e.g., (Durai et al., 2016; Malouff et al., 2011; Mazurek et al., 2019; Pattyn et al., 2016; Wallhausser-Franke et al., 2012; Ziai et al., 2017; Zirke et al., 2013)). The direction and causality of this link remain unclear, as pointed out in many previous studies, although individuals’ emotional states appear to be an important factor mediating the effects of tinnitus loudness on tinnitus-related distress (Probst et al., 2016a; Probst et al., 2016b; Schlee et al., 2016); anxiety, somatization, and in particular depression have also been identified as possible mediators of tinnitus-related distress (Bartels et al., 2010a; Bartels et al., 2010b; Trevis et al., 2016a; Trevis et al., 2016b).

The clinicians in our department noticed that the tinnitus patients seen since the hospital was reopened after COVID-19 had more emotional complaints than before. We thought that this might be related to the various pressures experienced by the patients during the pandemic event and the lockdown. Therefore, the COVID-19 pandemic and lockdown might provide a good opportunity to investigate whether anxiety impacts tinnitus as a promoting or enhancing factor. The present study explored whether anxiety was increased by the COVID-19 pandemic in subjects with tinnitus, and if so whether the increased anxiety affected the severity of tinnitus and the outcomes of tinnitus treatments.

## Methods

### Study Design

In this retrospective study, clinical data from outpatients visiting our department (the Hearing Center of Otolaryngology Department of the Sichuan Provincial People’s Hospital and Sichuan Academy of Medical Sciences, Chengdu, Sichuan, People’s Republic of China) were collected over the same periods, from March 1 to April 14, in both 2020 and 2019. This period in 2020 was the first 6 weeks of the reopening of our department to non-emergency visits after the nationwide lockdown for COVID-19 in China (from January 23 to February 29, 2020) that coincided with the deceleration phase of the pandemic and the resumption of economic activities. In this period, there were concerns about a resurgence of COVID-19 (Bedford et al., 2020).

The same protocol was followed for the treatment of patients during both years. On the initial visit, after collecting their history, every patient received a comprehensive audiological and psychological assessment. After the assessment, they were treated with one of three methods based on reported efficacy, financial cost, and the patient’s preference: sound therapy (ST) with educational counseling (EC) or relaxation therapy, sound amplification with EC and relaxation therapy, or EC and relaxation therapy without further treatment. Two months after the initial appointment, every participant was examined in a second assessment. Figure 1 shows a flowchart of the major procedures of this study. Although no procedure was experimental, we sought and received approval for the study from the Ethics Review Board of the Sichuan Provincial People’s Hospital and Sichuan Academy of Medical Sciences (permit number: 2020–355). This study was conducted according to the principles expressed in the Declaration of Helsinki (World Medical Association, 2018).

**Figure 1.**
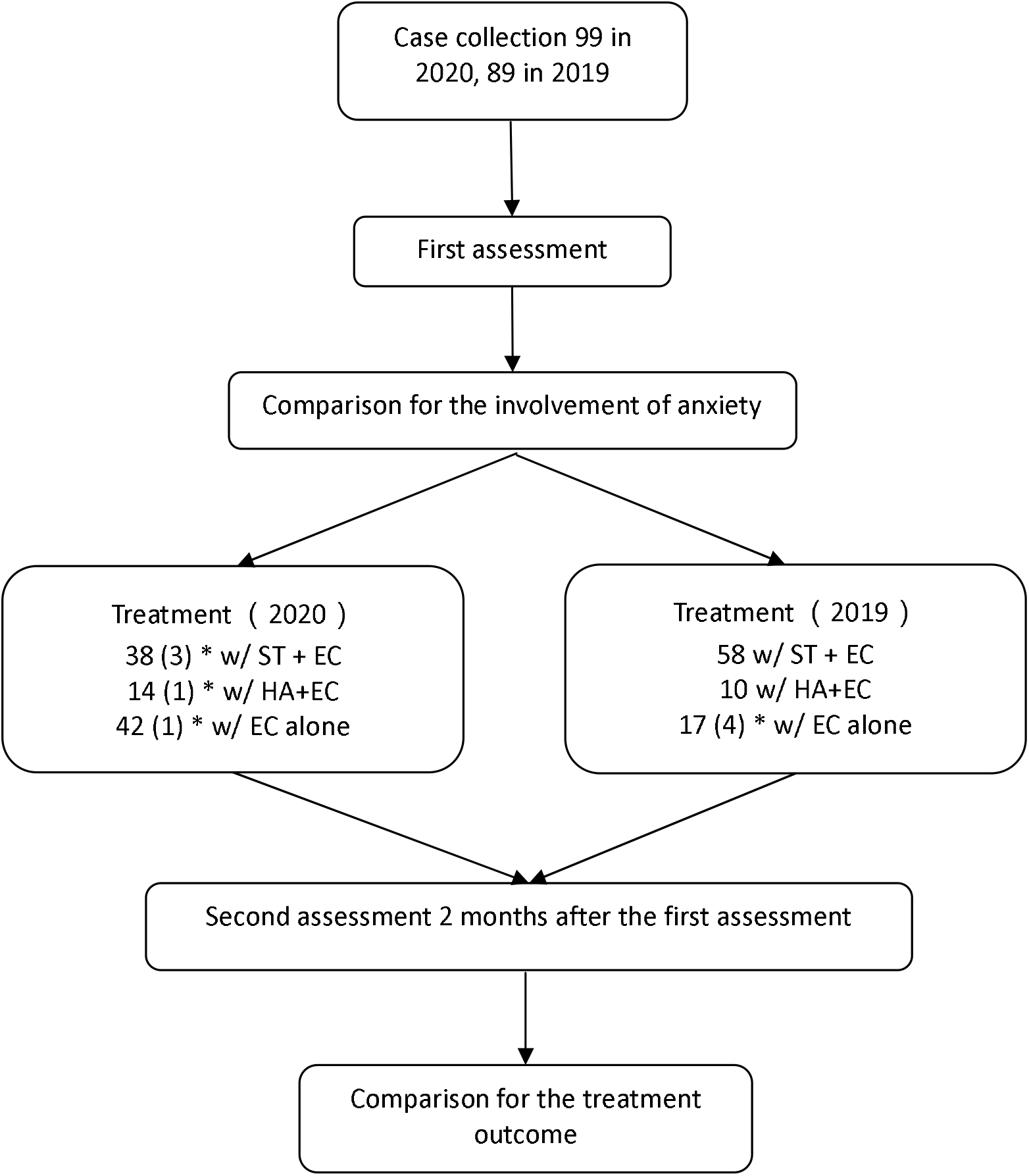
Flowchart of the major procedures in this study. *numbers in parentheses are those of cases that were lost to the study. THI: tinnitus handicap inventory, SAS: Zung’s Self-Rating Anxiety Sale, ST: sound therapy, HA: hearing aid, EC: educational counseling.

### Audiological Tests and Tinnitus Evaluation

The procedures for all tests were explained to the patients before they were conducted. All patients were examined using monocular otoscopy to identify any sign of blockage or inflammation in ear canals or perforation in the tympanic membrane. Tympanometry was tested at the most common 226 Hz probe tone, using an AT235 impedance meter (Interacoustics, Assens, Denmark); the type of tympanogram was determined for each ear (with type A as normal). Those who were abnormal in those tests were not included in this study.

The hearing status was tested with pure-tone audiometry (AC40, Interacoustics) in a soundproofed room. The air conduction threshold was examined for frequencies ranging from 250 Hz to 8 kHz using TDH 39 headphones (Telephonics, NY, USA) and bone conduction hearing was examined from 500 Hz to 4 kHz using a B-72 bone-conduction vibrator (Radioear, PA, USA), each in octave steps. The hearing thresholds were determined at each frequency using the standard Hughson–Westlake up–down procedure. Thresholds of 20 dB HL or lower were considered normal. The minimum masking level (MML) was tested in each ear with tinnitus, this test evaluates the maskability of tinnitus by external sounds. Broadband noise with a flat power spectrum was used for this evaluation, which was generated by a table-top sound generator (BTD01, BetterLife Medical Technology Co., Ltd., Jiangsu, China). To measure the MML, the level of the noise was gradually increased by the tester in 1 dB steps until the patient stated that the tinnitus had become nearly inaudible, then this level was recorded as the MML.

### Educational Counseling and Relaxation Therapy

The counseling was performed by the audiologists for each patient with tinnitus to acknowledge the patient’s suffering, and to help the patient understand tinnitus, demystify the condition, and correct any false preconceptions (duration 1 h) (Langguth, 2015). Relaxation therapy consisted of home-based exercises, such as listening to music, avoiding unnecessary tension, and tai chi (Arif et al., 2017; Tyler, 2014). Patients were advised to execute this for two sessions of 30 min per day over a period of 8 weeks.

### Sound Therapy

The first step of the ST was to identify the nature of the tinnitus in pitch and loudness. Pitch matching was conducted using the same sound generator (BTD01) as in the MML test to produce pure tones for tonal tinnitus or narrow-band noise for non-tonal tinnitus. The match was established by adjusting the central frequency and bandwidth, which could be changed from 100 Hz to 1 kHz, around the center frequency. In loudness matching, the matched tone or noise was presented continuously, and the level of the matching signal was adjusted from low to high until the tinnitus could hardly be heard. In this report, loudness matching results are presented in dB SL. Using the pitch and loudness matching data, a sound file was generated for each individual to produce a sound matching their tinnitus in frequency and level. This sound file was the uploaded to an ear level sound generator (BTM-N6, BetterLife Medical Technology Co., Ltd.) that was dispensed to the patient. The patients were instructed to listen to the sound file for 30 min each time, and to gradually increase from once to 3–6 times per day, every day, during the whole course of home-based therapy, which lasted for 2 months.

### Questionnaires

The tinnitus patients recruited in this study all completed two questionnaires at the initial visit and again during the follow-up, two months later. The Chinese version of the Tinnitus Handicap Inventory (THI) questionnaire was used in this study (Kam et al., 2009), consisting of 25 questions to assess the difficulty caused by tinnitus with respect to its functional, emotional, and catastrophic aspects (Meng et al., 2012; Newman et al., 1996).

A Chinese version of Zung’s Self-rating Anxiety Scale (SAS) questionnaire was used, which was adapted from a previous report (Gao et al., 2011; Zung, 1965). The raw scores were multiplied by 1.25 to generate the index scores (Zung, 1965). We used a value of 45 as the cut-off for anxiety, instead of 50, as reported in the most recent publication (Dunstan et al., 2020).

### Statistical Analyses

All parametric data are presented as mean ± standard deviation unless otherwise specified. When the parameters of participants were compared between two groups, the t-test was used or, if among multiple groups, analysis of variance (ANOVA) was used for continuous variables and the chi-square test for categorical variables, including sex, age, and site of tinnitus, and for risk factors among groups. Treatment outcomes were evaluated by comparing the scores of THI and SAS before and after the treatments, using a paired t-test or ANOVA. All analyses were performed using the SPSS 19.0.0 software at a significance level of 0.05. In figures, the significant level was indicated by the number of symbols (e.g., *), with 1, 2 or 3 representing p <0.05, 0.01, and 0.001 respectively.

## Results

A total of 99 cases were collected between March 1 and April 14, 2020, and 89 in the same period in 2019 (Figure 1). Table 1 compares the demographics and tinnitus characteristics between the subjects in the different years. The case load for tinnitus appeared to be higher in 2020 than in the same period in 2019 (99 vs. 89, or an increase of 11.2%). Such an increase could be largely attributed to the accumulation of cases when all the non-emergency visits were suspended during the lockdown between January and February 2020. The two groups of different years were matched by all clinical characteristics except the incidence of anxiety.

**Table 1.** Comparison of initial clinical characteristics of patients between 2020 and 2019

|  | March-April<br>2020 | March-April<br>2019 | p value |
| --- | --- | --- | --- |
| Sex (M:F) | 43:56 | 43:46 | .502 |
| Age (year old, M ± SD) | 50.8 ± 15.1 | 52.6 ± 14.7 | .487 |
| Educational background |  |  | .812 |
| Bachelor and superior | 54 | 47 |  |
| Inferior to bachelor | 45 | 42 |  |
| Duration (month) | 25 ± 53.6 | 31.3 ± 50.4 | .108 |
| Site |  |  | .177 |
| Bilateral | 36 | 41 |  |
| Unilateral | 63 | 48 |  |
| Anxiety involved/total# | 74/99 (74%) | 53/89 (59%) | .026 |
| Risk factors |  |  |  |
| Sensorineural hearing loss | 69 | 65 | .614 |
| Noise exposure | 1 | 0 | 1 |
| Hypertension | 3 | 6 | .179 |
| Hyperthyroidism | 1 | 0 | 1 |
| Head/neck trauma | 1 | 0 | 1 |

Chi-square test was used for the between-group comparison, sex, educational background, site, anxiety and the risk factor of sensorineural hearing loss using, t-test on age, Mann-Whitney Rank Sum Test on Duration, Fisher's exact test on the risk factors of noise exposure, hypertension, hyperthyroidism and head/neck trauma.

### The increase in Anxiety in 2020 and its impact on THI and MML

In the 2020 group, 74 out of 99 (or 74.7%) subjects had an SAS higher than 45 (the criterion for anxiety), which was significantly higher than that in the 2019 group (53/89, or 59%, χ^2^ = 4.938, p = 0.026). Overall, the SAS score in 2020 group was significantly higher than that of 2019 group (61.9 ± 11.9 in 2020 versus 49.1 ± 8.6 in 2019; U = 6867 via Mann-Whitney Rank Sum Test, p < 0.001, Figure 2A), which was fully due to the difference in the anxiety subgroups (68.0 ± 6.4 in 2020 vs. 54 ± 8 in 2019; U = 3550 via Mann-Whitney Rank Sum Test, p < 0.001, Figure 2A). Therefore, the higher SAS in 2020 was not simply due to the higher incidence of subjects with anxiety, but also the higher level of anxiety in the involved subjects.

**Figure 2.**
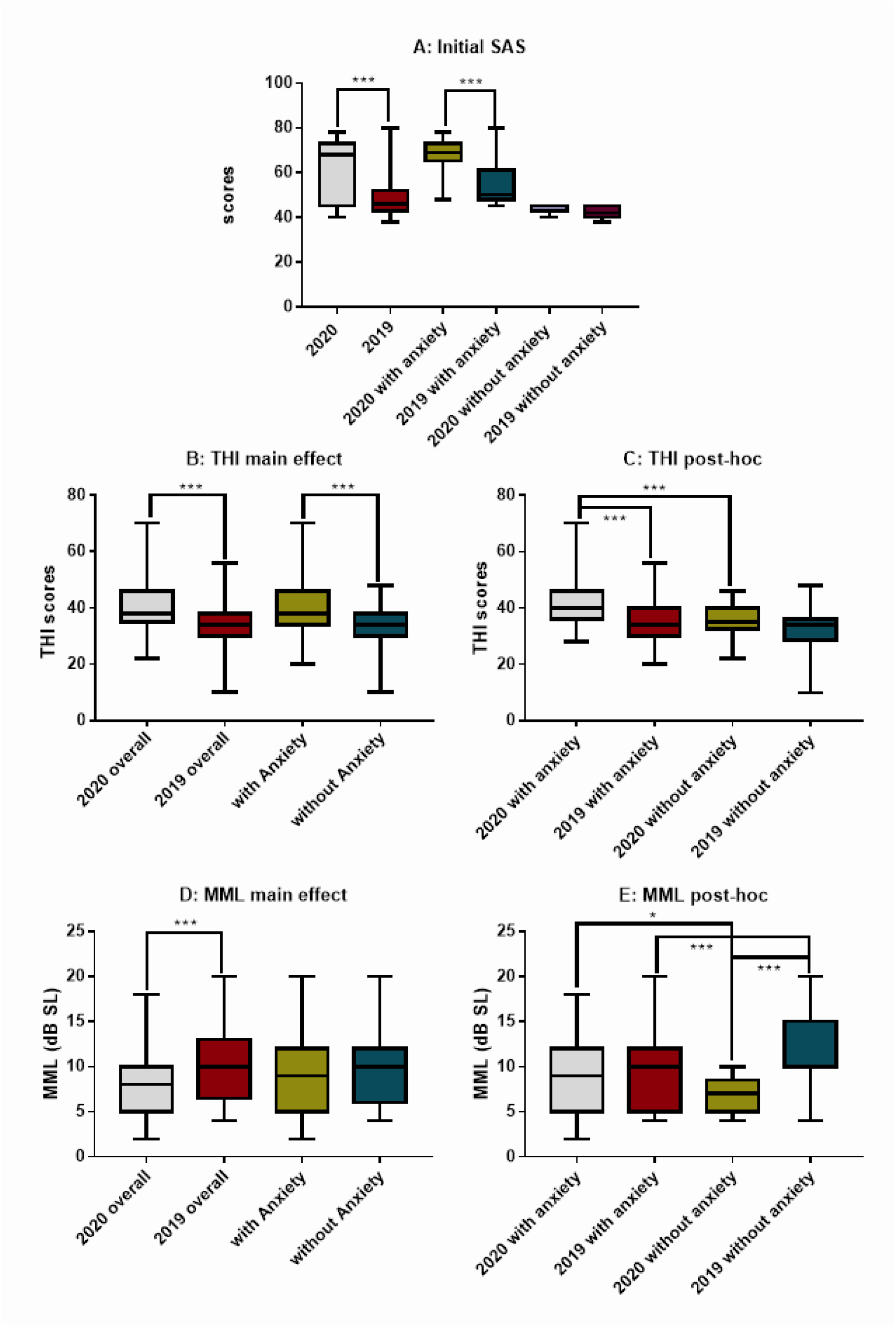
Comparisons of initial SAS, THI scores and MML between years and subjects with and without anxiety. A: SAS showing a significant difference between years and between the subgroups within the two years. B and D: The differences in THI and MML as the result of the two main factors—year and anxiety. C and E: *Post-hoc* comparison on THI and MML showing the difference within the factors of year and anxiety respectively. Within 2020, subjects with anxiety appeared to have a significantly higher THI and MML; no difference was seen in THI between anxiety and non-anxiety subgroups within 2019, while a higher MML was seen in non-anxiety subgroup within 2019. THI: tinnitus handicap inventory, SAS: Zung’s Self-Rating Anxiety Sale, MML: minimum masking levels.

The THI score in the 2020 group was 40.1 ± 6.9, which was significantly higher than that in the 2019 group (34 ± 8.3) as shown by the group effect in a two-way ANOVA against year group and anxiety (F_1,184_ = 16.278, p < 0.001). The ANOVA also demonstrated a significant effect of anxiety: 38.8 ± 8.6 for subjects with anxiety and 33.8 ± 7.5 for those without (F_1,184_ = 11.628, p < 0.001, Figure 2B). However, there was not a significant interaction between two factors (F_1,184_ = 2.3, p = 0.131). *Post-hoc* pairwise comparisons showed that the THI score of anxiety subgroup in 2020 was 41.7 ± 7.7, which was significantly higher than the corresponding subgroup in 2019 (34.8 ± 8.1; q = 6.904, p < 0.001), and that of non-anxiety subgroup in 2020 (35.6 ± 5; q = 4.766, p < 0.001, Figure 2C). Interestingly, the THI of non-anxiety subgroup in 2020 was (almost) same as that of the anxiety subgroup in 2019. However, there was no significant difference in THI score across the non-anxiety subgroups between years (Figure 2C).

The between-year difference in THI was further analyzed using a breakdown of the scores in the emotional, functional, and catastrophic questionnaire sections. A significant between-year difference was demonstrated in the emotional score (14.636 ± 3.7 in 2020 and 12.3 ± 3.3 in 2019; the Mann–Whitney rank-sum test, U = 5942.5, p < 0.001), in the functional score (18.515 ± 3.6 in 2020 and 15.5 ± 4.2 in 2019, U = 5211.5, p < 0.001) and in the catastrophic scores (7.0 ± 2.5 vs. 6.1 ± 2.6, U = 5173, p = 0.035). This result suggests that the higher THI in 2020 could be partially related to the increase in anxiety.

A two-way ANOVA similar to that for THI showed a significant year effect with subjects in 2020 had significantly lower MMLs (8.3 ± 3.5 dB SL) as compared to those in the 2019 group (10.4 ± 4.3 dB SL; F_1,184_ = 21.745, p < 0.001). However, the effect of anxiety was not significant (F_1,184_ = 0, p = 0.977; Figure 2D). The higher MML in 2019 could be largely attributed to the high MML in the non-anxiety subgroup this year as demonstrated by the *Post-hoc* pairwise test, which showed that the non-anxiety subgroups had a higher MML (11.7 ± 4.1 dB SL) in 2019 than the patients with anxiety in 2019 (9.5 ± 4.3 dB SL, q = 3.627, p < 0.001, Figure 2E). Within 2020, however, the anxiety subgroup had an MML of 8.9 ± 3.7 dB SL, which was slightly but significantly higher than the non-anxiety subgroup this year (6.7 ± 2.0 dB SL, q = 3.441, p = 0.015; Figure 2E). The result suggests that there is no clear indication whether anxiety played a role in the loudness of tinnitus.

Pearson correlation was conducted between SAS and THI and MML respectively in each year. In 2020, a weak positive correlation was seen between SAS and catastrophic THI (r = 0.319, p = 0.001), but not to another two subscales of THI. In this year there is also a moderate correlation between SAS and MML (r = 0.337, p < 0.001). In 2019, however, the significant correlation was seen in any pair of measurement (p > 0.05).

### Anxiety and Treatment Outcomes

The 94 patients in the 2020 group completed their face-to-face follow-up 2 months after the first assessment, while this number was 85 in the 2019 group (Figure 1). The numbers of patients who received ST with EC (STEC), hearing aids with EC (HAEC), or EC alone were 38, 14, and 42, respectively in the 2020 group, while the respective numbers were 58, 10, and 17 in the 2019 group. Due to the small sample sizes in patients receiving hearing aids in 2020, we only analyzed the treatment outcomes of STEC and EC alone. No between-year differences were seen in basic demographic features, risk factors and duration of tinnitus between the years in subjects treated with STEC (Table 2) and EC alone (Table 3). The incidence of anxiety in the patients receiving STEC was higher in the 2020 group (Table 2), but not such year difference was seen in patients received EC alone (Table 3).

**Table 2.** Between-year match in the demographic and selected clinic features in tinnitus patients treated with STEC

|  | May-June 2020 | May-June 2019 | *p*-value |
| --- | --- | --- | --- |
| Sex (M:F) | 16:22 | 30:28 | .356 |
| Age (year old, mean ± standard deviation) | 48.2 ± 15.7 | 50.2 ± 14.1 | .629 |
| Educational background |  |  | .507 |
| Bachelor and superior | 21 | 36 |  |
| Inferior to bachelor | 17 | 22 |  |
| Duration of tinnitus (month) | 25.5 ± 43.7 | 31.8 ± 54.3 | .428 |
| Site |  |  | .454 |
| Bilateral | 18 | 32 |  |
| Unilateral | 20 | 26 |  |
| Anxiety involved/total # | 29/38 (76%) | 32/58 (55%) | .035 |
| Risk factors |  |  |  |
| Sensorineural hearing loss | 24 | 40 | .555 |
| Noise exposure | 0 | 0 | \ |
| Hypertension | 1 | 6 | .396 |
| Hyperthyroidism | 0 | 0 | \ |
| Head/neck trauma | 0 | 0 | \ |

**Table 3.**
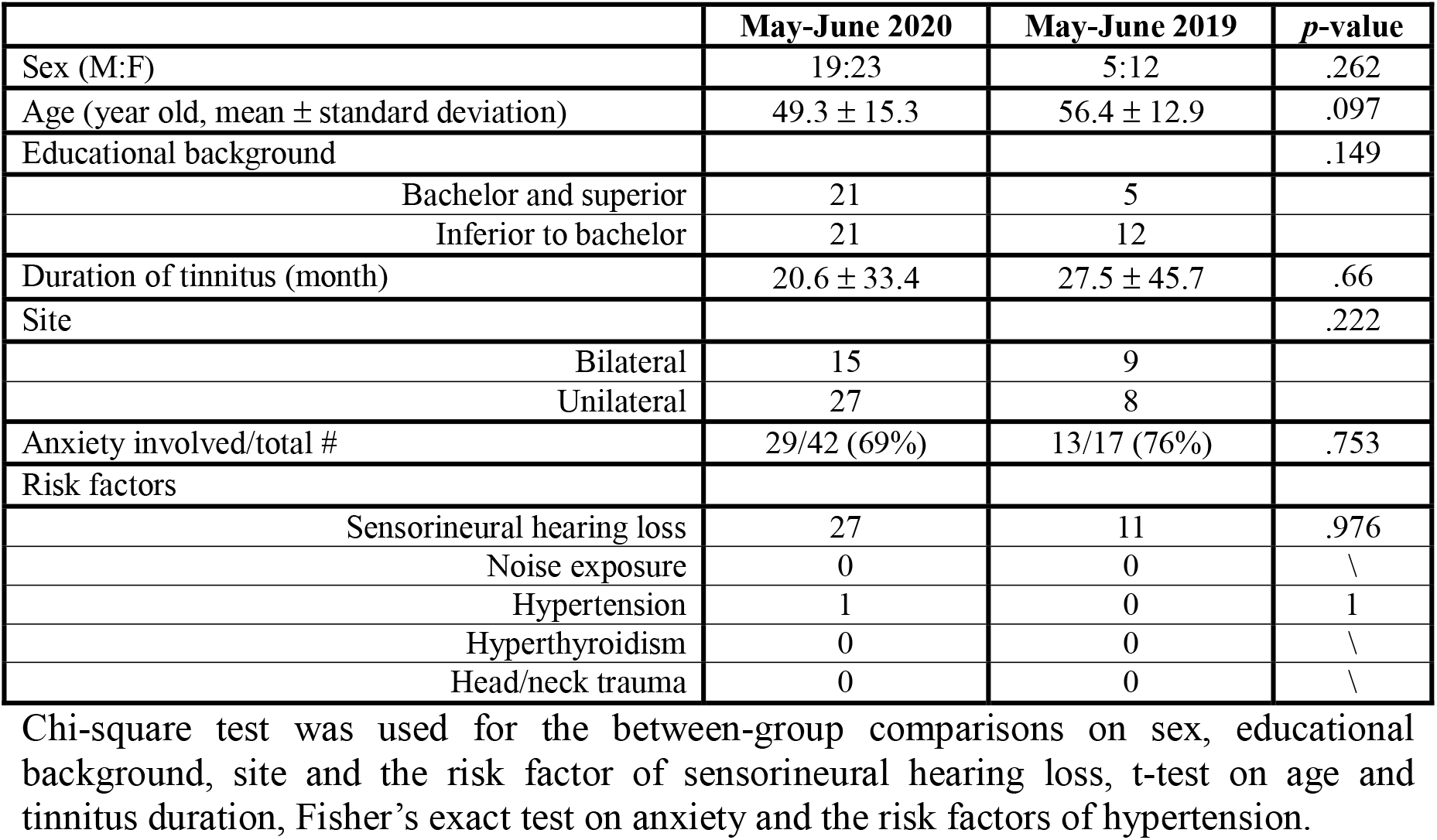
Between-year match in the demographic and selected clinic features in tinnitus patients treated with EC alone

### The effect of treatment on SAS

Figure 3 summarized the effect of the two treatments on SAS. In consistency with the data of whole sample (Figure 2A), the pre-treatment SAS was much higher in 2020 than in 2019 for the subjects treated with both STEC (Mann-Whitney Rank Sum Test, U = 411, p < 0.001, Figure 3A) and EC alone (U = 460.5, p = 0.031, Figure 3C). However, the effect of EC alone on SAS appeared to be qualitatively different from that of STEC in that the SAS was not decreased (improved) but increased in 2020 group after the treatment (Figure 3C), so that the post-treatment SAS in the 2020 group (63 ± 11) was even significantly higher than the before-treatment SAS in the 2019 group (52.9 ± 10, Mann–Whitney rank-sum test, U = 527, p < 0.004). This raised the question whether and how the number of subjects qualified as having anxiety changed after each treatment. Such changes were summarized in Table 4. In 2019, a large portion of subjects who had anxiety changed to non-anxiety status after either of the two treatments. In 2020, however, the number of cases with anxiety was increased, slightly after STEC, but largely after EC alone. In each method, there was a significant difference between years in the % change of cases with anxiety.

**Table 4.**
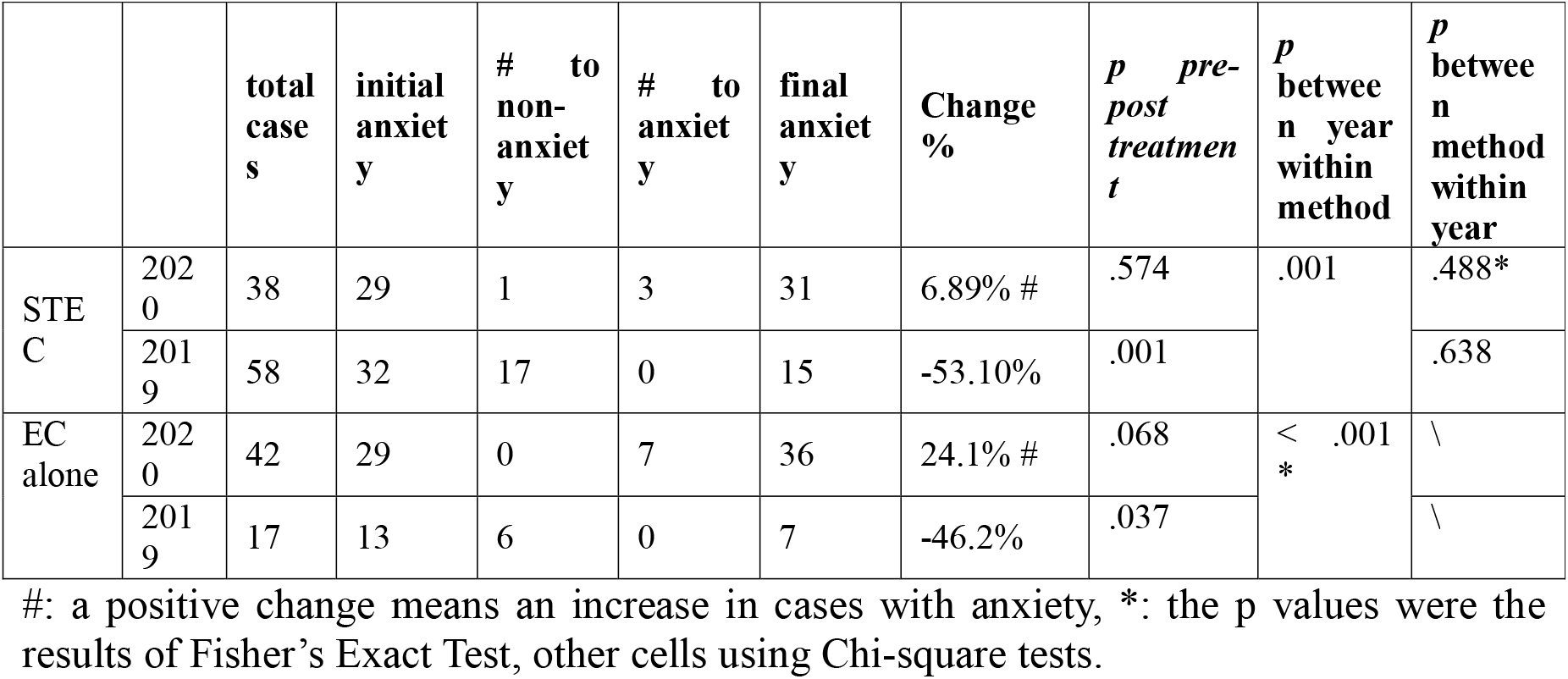
Changes of cases with anxiety after the treatments of STEC and EC alone.

|  |  | total<br>case<br>s | initial<br>anxiet<br>y | # to<br>non-<br>anxiet<br>y | # to<br>anxiet<br>y | final<br>anxiet<br>y | Change<br>% | *p* pre-post<br>treatmen<br>t | *p* betwee<br>n year<br>within<br>method | *p* betwee<br>n method<br>within<br>year |
| --- | --- | --- | --- | --- | --- | --- | --- | --- | --- | --- |
| STEC | 2020 | 38 | 29 | 1 | 3 | 31 | 6.89% # | .574 | .001 | .488* |
|  | 2019 | 58 | 32 | 17 | 0 | 15 | -53.10% | .001 |  | .638 |
| EC alone | 2020 | 42 | 29 | 0 | 7 | 36 | 24.1% # | .068 | < .001<br>* | \ |
|  | 2019 | 17 | 13 | 6 | 0 | 7 | -46.2% | .037 |  | \ |

#: a positive change means an increase in cases with anxiety, \*: the p values were the results of Fisher's Exact Test, other cells using Chi-square tests.

**Figure 3.**
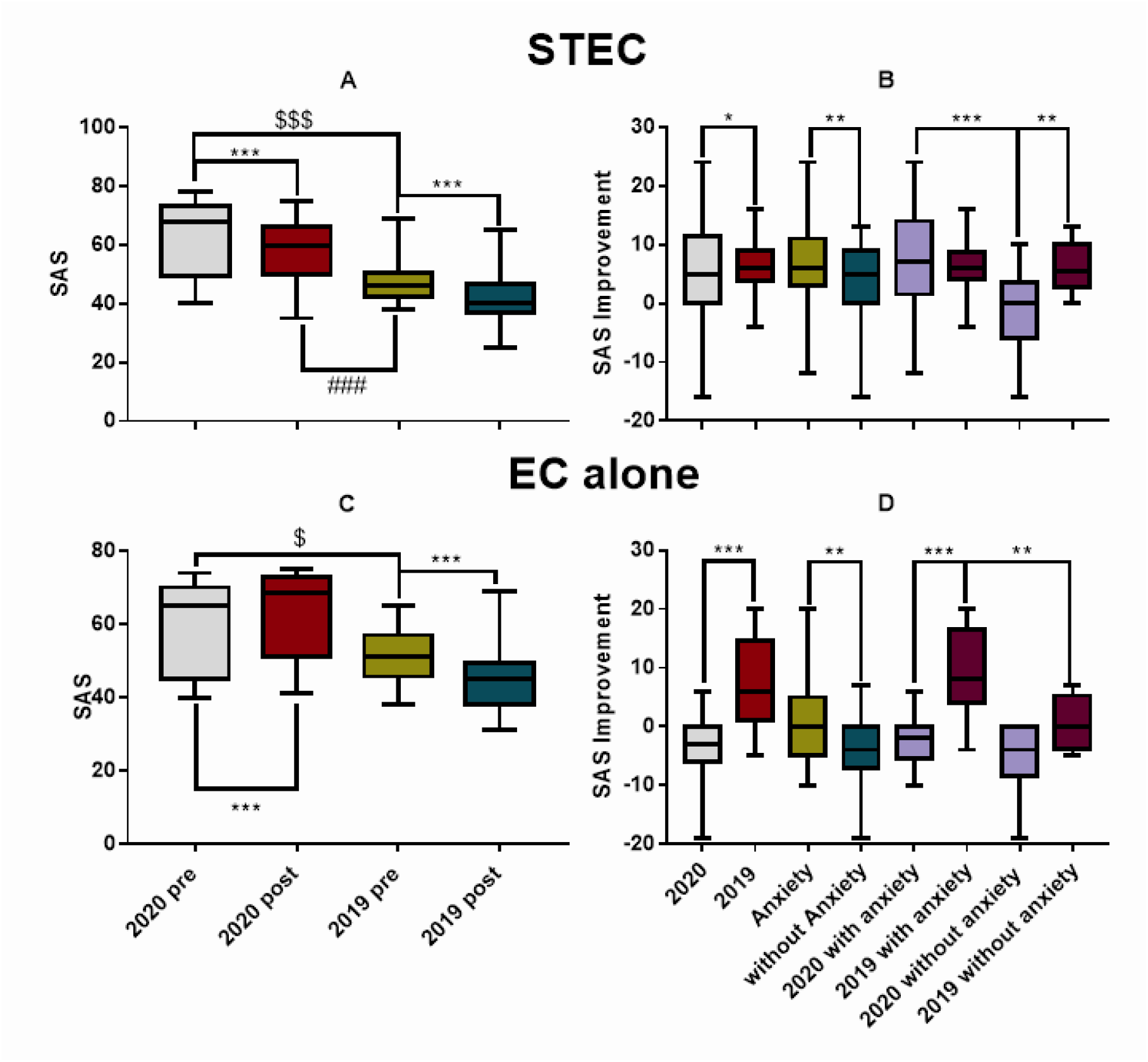
The SAS difference before and after the treatment of STEC (upper panels) and EC alone (the lower panels). A and C: The pre- and post-SAS. B and D: the pre-post difference of SAS score. STEC treatment reduced SAS in both years (A). However, EC alone did not improve SAS in 2020, instead the SAS was increased significantly in the 2^nd^ assessment (C). Correspondingly, STEC produced a slightly better improvement in SAS in 2019 than in 2020, but improvement by EC alone was much better in 2019 than in 2020, in which SAS was deteriorated. The number of symbols (*, $ or #) represents the level of significance, with 1, 2 or 3 symbols for p < 0.05, 0.01, or 0.001 respectively. STEC: sound therapy + educational counseling, EC: educational counseling.

The SAS was significantly reduced in both years after the STEC treatment (Mann-Whitney Rank Sum Test, p < 0.001). However, due to the large initial difference, the post-treatment SAS score in the 2020 group (58.0 ± 10.6) was still significantly higher than the pre-treatment SAS in the 2019 group (48.3 ± 8.5, Mann–Whitney rank-sum test, U = 534, p < 0.001). These results suggest that the anxiety associated with COVID-19 was not been fully counteracted by the treatment.

To further evaluate the effect of STEC on anxiety, a two-way ANOVA was performed on the pre-post SAS difference against the factor of year group and anxiety (Figure 3B). A significant year difference was seen since the SAS improvement appeared to be slightly but significantly smaller in 2020 (5.0 ± 8.6) than in 2019 (6.1 ± 3.8, F_1,92_ = 6.046, p = 0.016). Combined with the higher initial SAS in 2020, this implies that the higher initial anxiety in 2020 may have made the treatment less effective in reducing anxiety. However, this assumption is conflicted with the fact that the subjects with anxiety gained more reduction in SAS after STEC (6.6 ± 6.2 in the subjects with anxiety vs. 4.0 ± 5.9 in the non-anxiety subjects; effect of anxiety: F_1,92_ = 10.447, p = 0.002). Furthermore, the *post-hoc* test within 2020 revealed a larger SAS reduction (7.0 ± 8.0) in the anxiety subgroup this year than the non-anxiety subgroup in which the SAS was increased (negative improvement: -1.5 ± 7.2, *post-hoc* test within 2020, Tukey method; q = 5.364, p < 0.001). This result was in sharp contrast with the null difference in the SAS improvement between the anxiety subgroup (6.6 ± 6.2) and the non-anxiety subgroup (6.0 ± 3.5) in 2019 (Figure 3B).

A two-way ANOVA similar to the STEC was done for EC alone and showed a significant effect of year group: the pre-post difference in SAS in 2020 was negative (-3.4 ± 4.6, for an worse SAS) as compared with the large improvement in 2019 (7.1 ± 7.5; F_1,55_ = 26.022, p < 0.001). Since the initial SAS in the subgroup in 2020 receiving STEC was not significantly different from that in the subgroup receiving EC alone this year (63 ± 12 *versus* 59.5 ± 12.1; Mann–Whitney rank-sum test, U = 640.5, p = 0.129), the deteriorated SAS after EC alone suggests that the subjects in the EC subgroup in 2020 had experienced an increased stress after the first assessment, and the stress largely increased anxiety, which was not counteracted by the EC alone treatment. A significant effect of anxiety was also seen in subjects treated with EC alone: the SAS change after EC was 1 ± 7.4 in patients with anxiety before EC and -4 ± 6 in those without (F_1,55_ = 11.038, p = 0.002). There was no significant interaction between the two factors (F_1,55_ = 2.773, p = 0.102). The large deterioration in SAS in the non-anxiety subjects received EC is obviously due to such change in 2020 in which the SAS changes in the non-anxiety subjects was -5.4 ± 5.7, although this value was not significantly different from the change in non-anxiety subgroup in 2019 (0.5 ± 4.9; *post-hoc* test, q = 2.813, p = 0.052; Figure 3D). In both years, SAS improvement was smaller in the non-anxiety subgroups, and in 2020, SAS was deteriorated, instead of improved, in both anxiety and non-anxiety subgroups. In 2019, the SAS improvement in the anxiety subgroup 9.1 ± 7.1, which was significantly higher than the non-anxiety subgroup (0.5 ± 4.9) (*post hoc* test, via Tukey Method, q = 4.084, p = 0.006). In 2020, the SAS change in the anxiety subgroup was - 2.5 ± 3.8, and that in the non-anxiety subgroup was -5.4 ± 5.7. However, the difference was not significant (post hoc test, Tukey method, q = 2.324, p = 0.106). To further evaluate the impact of anxiety on clinic features of tinnitus, Pearson product moment correlation was calculated between the initial SAS score and the changes after the treatment. There was a moderate, positive, linear relationship between the initial SAS score and the change in patients receiving STEC in 2020 (r = 0.511, p = 0.001), but no significant correlation was found in 2019 (Figure 4A). In addition, a moderate and positive linear relationship was also seen between the initial SAS score and the change in patients receiving EC alone in 2020 (r = 0.413, p = 0.006; Figure 4B) but not in 2019 (r = 0.488, p = 0.071). These results suggest that the treatment was more effective for mitigating anxiety in subjects with higher SAS scores in 2020, which was associated with the COVID-19 pandemic.

**Figure 4.**
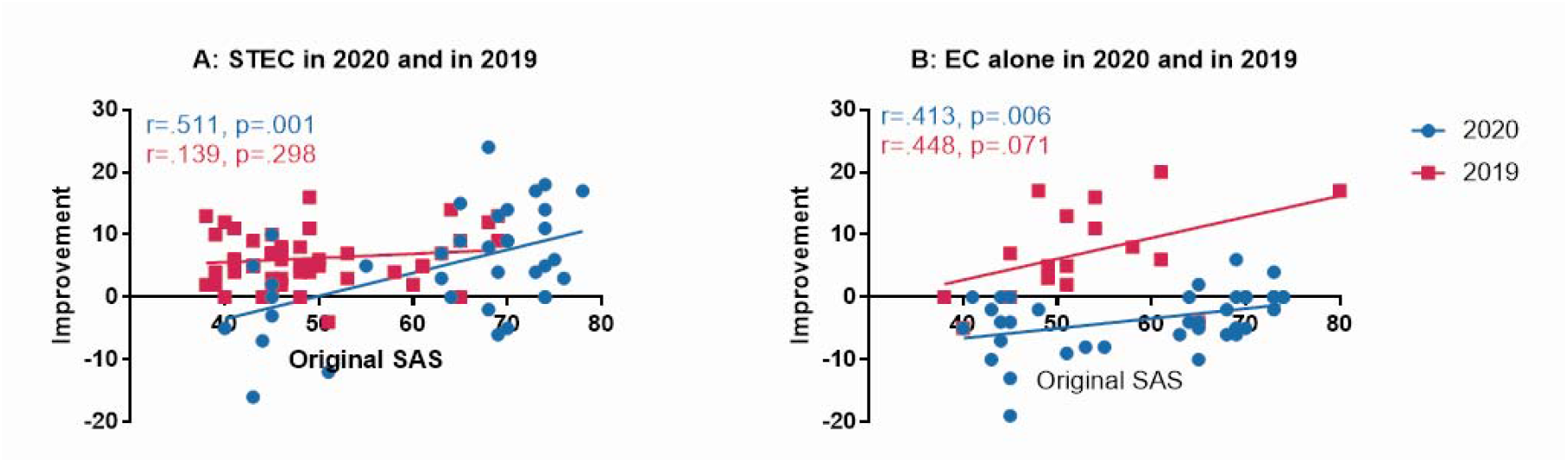
Correlations between the initial SAS score and the improvement in SAS score. A: The correlation of STEC by year. B: The correlation of EC alone by year. Significant, moderate correlations were seen in STEC group and EC alone groups in 2020 in which the average initial SAS scores were much higher. SAS: Zung’s Self-rating Anxiety Scale, ST: sound therapy, EC: educational counseling

### The effect of treatments on THI and MML

The effect of the treatments was first examined by self-reported improvement (reduction) of tinnitus loudness. As expected, the case number and rate reporting an improvement were higher in subjects treated with STEC than in those with EC alone in both years. More importantly, the case number with improvement was significantly lower in 2020 group than in 2019 in subjects treated with both methods (Table 5). However, there were no significant differences in the case rate reporting an improvement between subjects with and without anxiety (data not shown).

**Table 5.** Self-reported improvement of tinnitus loudness in the Follow-ups of treatment groups between years

|  | STEC group | EC alone group | p between methods |
| --- | --- | --- | --- |
| 2020 | 27/38 (71%) | 8/42 (19%) | < 0.001* |
| 2019 | 51/58 (88%) | 9/17 (53%) | 0.004** |
| p between year | .038* | .024** |  |

\*: chi-square test, \*\*: Fisher's Exact Test

STEC significantly reduced the THI scores in both 2020 group from 40.7 ± 6.7 to 37.7 ± 8.0 (via paired t-tests, t_0.05/37_ = 3.253, p = 0.002) and 2019 group from 32.7 ± 8.3 to 28.7 ± 7.6 (via Wilcoxon Signed Rank Test, W = -1590, p < 0.001) as shown in Figure 5A. Figure 5B summarized the result of a two-way ANOVA on the improvement of THI (the pre-THI minus post-THI) by STEC against the factor of year group and anxiety. There was no significant effect for both factors (year effect: F_1,92_ = 2.104, p = 0.15; anxiety effect: F_1,92_ = 0.09, p = 0.759).

**Figure 5.**
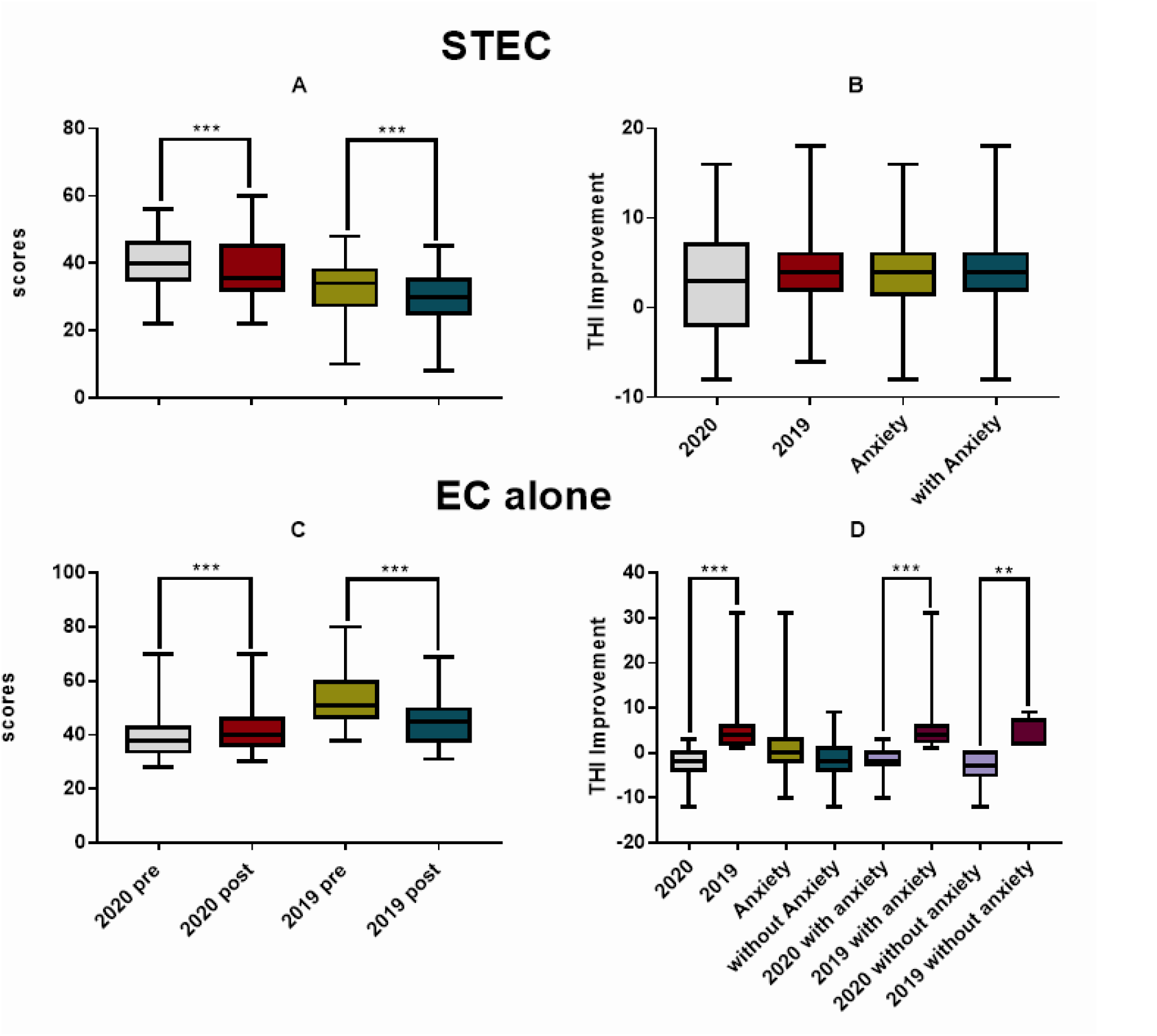
The difference in THI before and after the treatment of STEC (upper panels) and EC alone (the lower panels). A and C: The pre- and post-THI scores. B and D: the pre-post difference of THI score. STEC resulted in a significant THI reduction in both years (A), but there was no significant difference in the amount of reduction between years and between subjects with and without anxiety (B). EC alone reduced THI in 2019, but opposite in 2020 (C and D). The THI got deteriorated in 2020 and worse than 2019 in both subgroups with and without anxiety (D). Therefore, within subjects with or without anxiety, the treatment resulted in a better THI in year 2019. STEC: sound therapy + educational counseling, EC: educational counseling.

Surprisingly, the THI scores in 2020 rose from 39.8 ± 8.9 to 42.1 ± 9.1 after EC alone treatment (Wilcoxon Signed Rank Test, W = 426, p < 0.001), while an improvement was seen in 2019 from 35.7 ± 5.2 to 30.2 ± 6.3 (Wilcoxon Signed Rank Test, W = -153, p < 0.001, Figure 5C). Therefore, the change in THI by EC alone was - 2.2 ± 2.9 in 2020, but 5.4 ± 6.9 in 2019, as shown by the significant year effect in the two-way ANOVA (F_1,55_ = 25.73, p < 0.001). In this ANOVA, the effect of anxiety was not significant (Figure 5D). Correspondingly, the between-year difference in THI improvement was larger in anxiety subjects than non-anxiety ones (*post hoc* tests, q = 7.323, p < 0.001 in anxiety between year and q = 4.031 p =0.006 in non-anxiety between year).

Correlation analysis showed a moderate and positive linear relationship between the improvements of THI in the emotional subscale and the SAS improvement in the subjects treated with STEC in both 2020 (r = 0.506, p = 0.001) and 2019 (r = 0.623, p < 0.001; Figure 6A). In subjects treated with EC alone, significant correlation was seen only in 2019 group (r = 0.536, p < 0.026) but not in 2020 group (Figure 6B).

**Figure 6.**
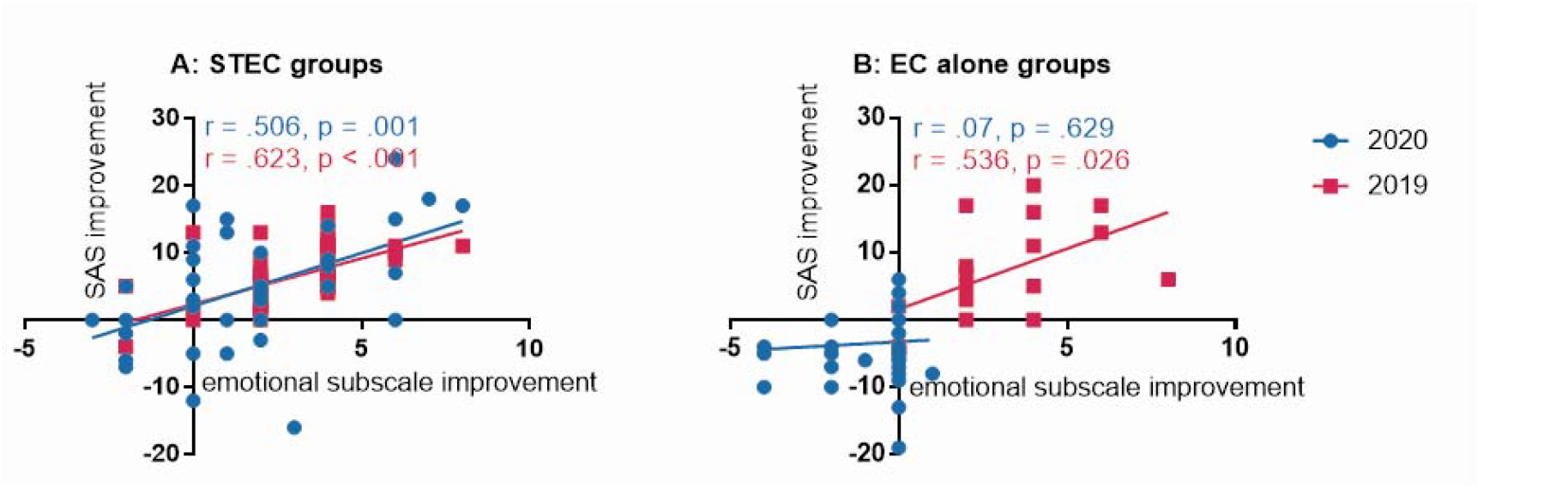
Correlations between the improvements of emotional section in THI and SAS in 2020 and 2019. A: The correlation in ST-EC group. B: The correlation in EC alone group. Significant, moderate correlations were seen in ST with EC group in both years and EC alone groups in 2019. SAS: Zung’s Self-rating Anxiety Scale, ST: sound therapy, EC: educational counseling.

MML was reduced by STEC in 2020 group (from 9 ± 4.4 dB SL to 7.3 ± 4.2 dB SL; Wilcoxon Signed Rank Test, W = -391, p = 0.003) and 2019 group (from 10.0 ± 3.8 dB SL to 7.9 ± 3.8 dB SL; W = -1525, p < 0.001; Figure 7A). The improvement (2.1 ± 1.7 dB) was slightly higher in 2019 than in 2020 (1.6 ± 2.7 dB), but the difference was not statistically significant as shown by the main effect of year in a two-way ANOVA (F_1,92_ = 1.513, p = 0.222, Figure 7B). Neither a significant effect of anxiety was seen in this ANOVA (F_1,92_ = 0.006, p = 0.935).

**Figure 7.**
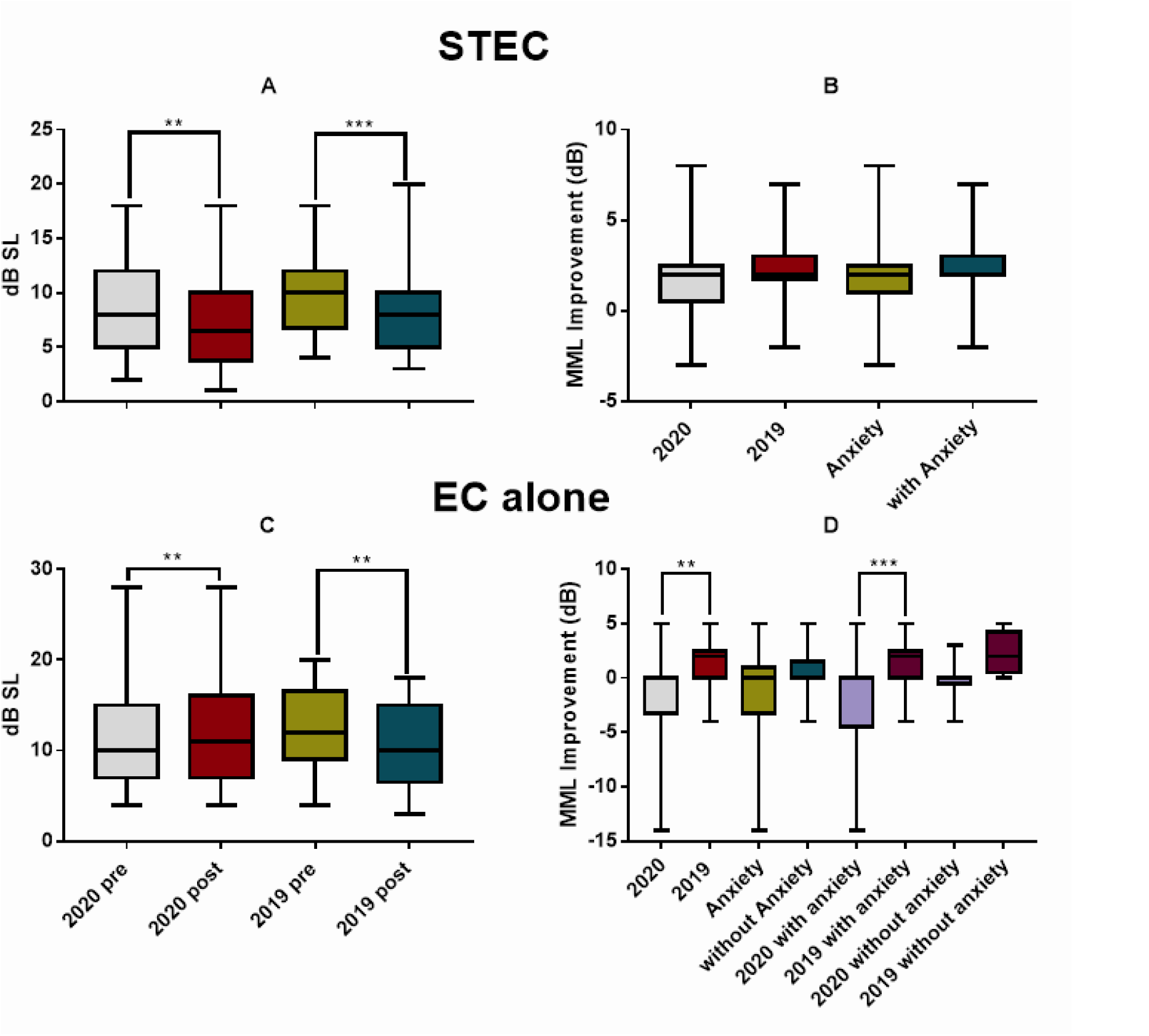
The changes in MML between the two assessments before and after the treatment of STEC (upper panels) and EC alone (the lower panels). A and C: The pre- and post-MML. B and D: the pre-post difference of MML. The MML got deteriorated in 2020 and worse than 2019 in the subgroup with anxiety (D). Significance: **p < 0.01, ***p < 0.001 in ANOVA. STEC: sound therapy + educational counseling, EC: educational counseling.

Like THI, EC alone treatment in 2020 did not reduced MML, but increased it from 10.9 ± 4.9 dB SL to 12.4 ± 5.8 dB SL (Wilcoxon Signed Rank Test, W = 172, p = 0.003), yielding an increase of 1.5 ± 3.1 (Figure 7D). This was opposite to the decrease in MML from 12.4 ± 4.9 dB SL to 10.3 ± 4.7 in 2019 (Wilcoxon Signed Rank Test, W = -91, p < 0.001; Figure 7C). Correspondingly, a significant year effect was seen in a two-way ANOVA (F_1,55_ = 10.036, p = 0.003), which did not show a significant effect of anxiety (F_1,55_ = 1.944, p = 0.169). However, the year difference was mainly due to the between-year difference in the anxiety subjects in the *post-hoc* test (Tukey method, q = 5.24, p < 0.001), since no significant difference was seen in non-anxiety subjects between years (q = 2.129, p > 0.05, Figure 7D). Moreover, correlation analyses did not show any significant correlation between initial SAS and the change of MML after both treatment in each of the two years. Those results suggest that high anxiety in 2020 made EC alone treatment ineffective in mitigating loudness of tinnitus. The overall correlations between SAS improvements and THI (with subscale THI), MML improvements by two treatment methods in two years were seen in Table 6.

**Table 6.** Correlation between SAS improvements and those in THI and MML

|  | **r** | **p-value** | **r** | **p-value** |
| --- | --- | --- | --- | --- |
| Target | (A) STEC in 2020 |  | (B) STEC in 2019 |  |
| THI Total | 0.459 | 0.003 | 0.193 | 0.146 |
| THI Functional | 0.17 | 0.307 | -0.379 | 0.003 |
| THI Emotional | 0.506 | 0.001 | 0.623 | < 0.001 |
| THI Catastrophic | 0.313 | 0.055 | 0.149 | 0.265 |
| MML | 0.134 | 0.424 | 0.143 | 0.286 |
|  | (C) EC in 2020 |  | (D) EC in 2019 |  |
| THI Total | 0.3 | 0.053 | -0.008 | 0.975 |
| THI Functional | 0.313 | 0.04 | -0.347 | 0.172 |
| THI Emotional | 0.07 | 0.629 | 0.536 | 0.026 |
| THI Catastrophic | 0.112 | 0.481 | -0.04 | 0.856 |
| MML | 0.117 | 0.461 | 0.222 | 0.392 |

## Discussion

Several interesting findings were seen in this retrospective study. (1) We demonstrated a significantly increased anxiety in the tinnitus subjects seen in 2020 in terms of the incidence of subjects with anxiety (Table 1) and the averaged SAS (Figure 2A). Based upon the significant between-year difference, this increase in anxiety is clearly associated with COVID-19 pandemic. (2) The high SAS was associated with a high THI score, especially in the emotional subscale in 2020 as compared with the values of 2019 (Figure 2B and 2C), suggesting that the increased psychological stress in 2020 does enhance tinnitus. (3) However, the increased anxiety was not clearly linked to measure of tinnitus loudness by MML (Figure 2D and 2E). (4) Overall, the treatments of both STEC and EC alone were less effective in 2020 in anxiety reduction (Table 4 and Figure 3B and 3D) and in the self-reported mitigation of tinnitus (Table 5). In fact, the anxiety was even worse after the treatment in 2020, especially in those who received EC alone. This suggested that an increased stress was experienced by the subjects in 2020 group after the first assessment, which could not be counteracted by the therapy. (5) There was no significant difference between years for the reduction of tinnitus severity as measured by THI and MML by STEC (Figure 4B and 5B). (6) However, the treatment of EC alone was much less effective in reducing THI and MML, and in 2020 it resulted in a deterioration increase in anxiety (Table 4, Figures 3D, 4D), THI (Figure 5D) and in MML (Figure 7D). Since EC alone did show benefit in 2019, the deterioration in 2020 suggests that the anxiety in 2020 largely enhanced tinnitus, and made it difficult to be managed.

There is no doubt that a significant psychological stress was developed because of the COVID-19 pandemic. Many recently published articles have revealed the high prevalence of anxiety across China during the COVID-19 pandemic, from 28.8% to 35.1% (Huang et al., 2020; Wang et al., 2020a), as compared to the previously reported prevalence of 5.6% and 7.6% for the years of 2009 and 2019, respectively (Huang et al., 2019; Phillips et al., 2009). A cross-sectional survey, using the same anxiety questionnaire as adapted in the present study, reported an average SAS score of 45.89 ± 1.1 among front-line clinical staff during the pandemic (Wu et al., 2020). This value was located between the scores for our subjects with and without anxiety (68.0 ± 6.0 vs. 43.9 ± 1.5), and lower than the average for all subjects in the 2020 group (61.9 ± 11.9). This implies that our tinnitus patients seen in 2020 have experienced extremely high psychological pressure, even higher than those medical doctors who were in the most challenging job during the pandemic. The number of tinnitus subjects seen in the 6-week period in 2020 was higher than that last year. However, this increase may be largely attributable to the accumulation of patients during the hospital closure in the national lockdown.

The association between tinnitus and anxiety has been investigated in many previous studies and has been well reviewed (Durai et al., 2016; Malouff et al., 2011; Mazurek et al., 2019; Pattyn et al., 2016; Wallhausser-Franke et al., 2012; Ziai et al., 2017; Zirke et al., 2013). However, no information is available on the direction and causality between the two ends of the link (Danioth et al., 2020; Lugo et al., 2020; Mazurek et al., 2019; Park et al., 2019b; Wallhausser-Franke et al., 2012), although many studies have implied that psychological states, such as those related to common stressors, influence perception of, or coping with tinnitus (Lazarus, 1993; Lazarus, 2000). In this regard, two related systems are involved in tinnitus: (1) the brain regions along the hypothalamic–pituitary– adrenal axis (see reviews (Mazurek et al., 2019; Ziai et al., 2017)), which is the main neuroendocrine system involved in stress response, and (2) the limbic system including the hippocampus and amygdala, which regulates the perception of tinnitus and the adaptation (thereby, the ability to cope with stress) (Chen et al., 2017; Kapolowicz et al., 2019; Leaver et al., 2016; Lockwood et al., 1998; Raghavan et al., 2016; Zhang et al., 2015). While the data from the previous studies have indicated the possible role of emotional factors in tinnitus via those systems, the relationship was mostly investigated in animal models, or in cross-sectional comparisons across subjects with different levels of tinnitus and those without, with focus on establishing the connection, rather than on the directional nature of the link.

The COVID-19 pandemic provides a good opportunity to investigate whether stress or anxiety could enhance tinnitus as a causative or promotive factor, by clearing some clouds. For example, in many of the previous studies, the effect of anxiety on tinnitus were evaluated in a special population, such as those in veterans (Hu et al., 2015), in elderly (Danioth et al., 2020), in those with headache (Lugo et al., 2020), and those with sleeping disorders (Xu et al., 2016a). In other extreme, the link was investigated in cross-sectional studies in which the anxiety cases of different causes was included (Park et al., 2019a). Moreover, the anxiety has been evaluated with many different methods, including Hospital Anxiety and Depression Scale (McKenna et al., 2017), Beck Anxiety Index (Mahboubi et al., 2017), as well as SAS (Xu et al., 2016b). All those variations make it difficult to generalize a finding, if reported, for the directional nature of the link between anxiety and tinnitus. Although large variation existed across different individuals in relationship to their jobs and financial situations, as well as their closeness to COVID-19 patients, the stress factor associated with this study was much more homogeneous than those that had been examined in previous studies. Moreover, it has been shared by general population rather than impacting on small groups. In addition, the same methodologies were used over the two years, which ensured a valid comparison for verifying the impact of anxiety associated with COVID-10. We therefore think that the between-year differences in the tinnitus clinic afforded a good chance to verify whether anxiety plays a causative or promotive role for tinnitus.

In the present study, at least three lines of evidence pinpointed the causative/promotive role of anxiety on tinnitus. Firstly, the high anxiety (in both the case% and SAS) was associated with the higher THI in all three subscales in 2020. Secondly, the high anxiety reduced the effectiveness of the tinnitus treatment in 2020 as compared with 2019 result, in the change of SAS (Figure 3B and 3D), the case% of subjects with anxiety (Table 4), self-reported improvement in tinnitus loudness (Table 5) and THI (Figure 4D).

The results in Table 4 indicate a sharp contrast in the changes of cases with anxiety after the treatments between years: an increase of 6.89% by STEC in 2020 versus a decrease of 53.1% in 2019, an increase of 24.1% by EC alone in 2020 versus a decline of 46.2% in 2019. The between-year differences indicates that the higher-level stress in 2020 affected the efficacy of the two treatments in mitigating anxiety. Furthermore, the self-reported improvement in tinnitus loudness (Table 5) was also significantly less in 2020 in both treatments. Thirdly, the promoting/enhancing effect of anxiety on tinnitus was indicated by the significant difference in the treatment effectiveness between STEC and EC alone. To evaluate the full impact of the stress on tinnitus, an untreated control group would be ideally used. Unfortunately, we do not have such control. However, the EC alone treatment was given for only one time of 30-60 minutes session over the whole 2 months. This was not a comprehensive therapy by any means. Therefore, the EC subgroup could be used as a virtually no-treatment control, although this method exerted a “better than nothing” effect in 2019. We found a significant increase in SAS in the subjects treated with EC alone in 2020, while a reduction in SAS in the STEC subgroup. Since the initial SAS was not different between the subjects treated with the different methods, this difference suggests that there was an increased or accumulated anxiety during the two months after the first assessment in 2020, which could not be counteracted by EC alone treatment. There were no significant between-year differences in the change of THI and MML by STEC. However, the THI and MML got worse in 2020 EC alone subgroup in association with a large increase in SAS, while the same treatment somehow improved both THI and MML in 2019. These results suggest that the increased stress, if not treated effectively, have significantly enhanced the tinnitus in 2020.

EC is a psychological treatment that was often recommended in combination with other treatments, like sound therapy or hearing aid fitting (Brennan-Jones et al., 2020; Jastreboff et al., 2000). However, different effectiveness of EC alone was also reported in some studies. For instance, an early study reported a successful ratio of 18% in tinnitus release (Jastreboff et al., 1996); while another study reported a significant THI reduction from 46.11 ± 22.74 to 31.94 ± 20.41 (Liu et al., 2018). In the present study, the THI was reduced by 5.4 ± 6.9 after EC alone treatment in 2019. This result demonstrates the effectiveness of our EC treatment, while the quantitative difference between our data and others may reflect the detail difference in EC procedures and other factors such as subject variables. Anyway, the EC alone treatment reduced SAS (Figure 3D), THI (Figure 5D) and MML (Figure 7D) in 2019. However, the change of SAS, THI and MML occurred in the opposite direction after the EC in 2020. The between-year difference validates the use of EC alone as a virtual control because it is obvious that this treatment was not sufficient to counteract the effect of anxiety.

### Limitations

There were several limitations to our study. Firstly, this was a retrospective study in which only the SAS was used to evaluate anxiety. This makes it difficult to compare our study with previous ones. Secondly, STEC was compared with EC alone without the use of wait-list control, making it difficult to fully evaluate the impact of anxiety on tinnitus. Thirdly, more patients in 2020 selected EC alone treatment probably due to the financial constraints, which may have produced some bias in comparison with 2019 subgroup. Last but not least, the overall sample size in the present study was small as the data were collected only from one hospital within a limited period. Although the data and conclusion are solid in the present study, further investigation would be helpful to verify the conclusion with a larger sample.

Currently, the link between anxiety and tinnitus was more evaluated in the direction of how tinnitus, as a stressor, can interact with (pre-existing) psychological disorders and change the subjects responses to them (Kroner-Herwig et al., 2006), but was not emphasized on the direction whether other stressors would enhance tinnitus. This has been reflected in evaluation tools. For example, the THI questions for the emotional subscale (e.g., Question 22: Does your tinnitus make you feel anxious) obviously ask the impact of tinnitus on emotion, but there is no question asking whether a stressor changes the severity of tinnitus (Newman et al., 2008). This bias appears to be a limitation for investigating the causative role of anxiety or stressor on tinnitus, and is likely one of the reasons why there was only a week correlation between the large increase in SAS in 2020 and the THI in the initial assessment. In future investigation, THI questionnaire should be revised accordingly.

## Conclusion

A substantial increase in anxiety was seen in tinnitus subjects in 2020 in association with COVID-19 pandemic and was evident as a promoting factor to tinnitus. The increase in SAS was associated with a smaller increase of THI in 2020, but not by the difference in MML. However, the difference in treatment effect between STEC and EC alone suggested that, the tinnitus severity was increased (in both THI and MML) when it was not comprehensively treated (such as by EC alone). Therefore, the present study provided clear evidence for the promoting effect of anxiety on tinnitus.

## Data Availability

All relevant data are within the paper.

## Acknowledgement

The authors acknowledge the colleagues for participating in this study. We are deeply indebted to the families who participated in the study.

